# Investigating markers of pulmonary aspiration in bronchoalveolar lavage from children with severe neurodisability

**DOI:** 10.1101/2023.10.03.23295956

**Authors:** Rosemary E Maher, Ruth E Trinick, Mark Dalzell, Robert J Beynon, Paul S McNamara

**Author notes:** Corresponding author **Corresponding author:** Prof Paul McNamara, Department of Child Health (University of Liverpool), Institute in the Park, Alder Hey Children’s Hospital, Liverpool, United Kingdom, L12 2AP, United Kingdom. Joint first authors (Rosemary E Maher, Ruth E Trinick).

## Abstract

Respiratory exacerbations are a frequent cause of hospitalisation in children with severe neurodisability (ND). Direct aspiration of food/saliva, reflux aspiration of gastric contents or a combination of both is thought to be a common cause of respiratory symptoms and disease, particularly when this occurs ‘silently’. A number of aspiration biomarkers, including bile acids and pepsin, have been proposed, however, no gold-standard diagnostic tests are currently available. In children with severe ND at high risk of both direct and reflux aspiration, we analysed lower airway samples for saliva- and/or gastric-specific proteins with biomarker potential.

## Introduction

Respiratory exacerbations are a frequent cause of hospitalisation in children with severe neurodisability (ND). Direct aspiration of food/saliva, reflux aspiration of gastric contents or a combination of both is thought to be a common cause of respiratory symptoms and disease, particularly when this occurs ‘silently’.^1^ A number of aspiration biomarkers, including bile acids and pepsin, have been proposed, however, no gold-standard diagnostic tests are currently available.^2^ In children with severe ND at high risk of both direct and reflux aspiration, we analysed lower airway samples for saliva- and/or gastric-specific proteins with biomarker potential.

## Methods

Non-bronchoscopic bronchoalveolar lavage fluid (BALF) was collected prior to elective surgery from healthy control children (*Control*, n=13), and children with severe non-ambulant ND (*Elective-ND*, n=9) considered to be otherwise well by both parents, anaesthetists, and surgeons. BALF was also collected from severe non-ambulant children with ND intubated on PICU with respiratory exacerbations (*PICU-ND*, n=16). Four saliva and two gastric fluid samples were obtained from healthy adult controls.

Samples were processed, and proteomic analysis of saliva, gastric fluid and BALF undertaken as previously described.^3,4^ Highly expressed proteins in saliva and gastric fluid were identified through the Human Protein Atlas (https://www.proteinatlas.org/) and literature review. Only candidates with >1 unique peptide identification were used to focus a search in all BALF samples.

## Results

In total, 383 proteins were identified in saliva. Of these, only two of 15 saliva specific proteins were identified in BALF. Alpha-amylase-1, the most abundant salivary protein, was identified in saliva by 22 unique peptides and in BALF by 19 unique peptides. Its label-free abundance profile was highest in the elective-ND group and lowest in the PICU-ND group (Mann Whitney U p-value=0.00038, Figure 1). Lipocalin-1, a cysteine proteinase inhibitor also known as von Ebner’s gland protein, was identified in saliva by 6 unique peptides and in BALF by 9 unique peptides. Its abundance in BALF was similar to alpha-amylase-1, i.e., higher in elective-ND relative to PICU-ND and control groups (p-value=0.00012, Figure 1).

**Figure 1.**
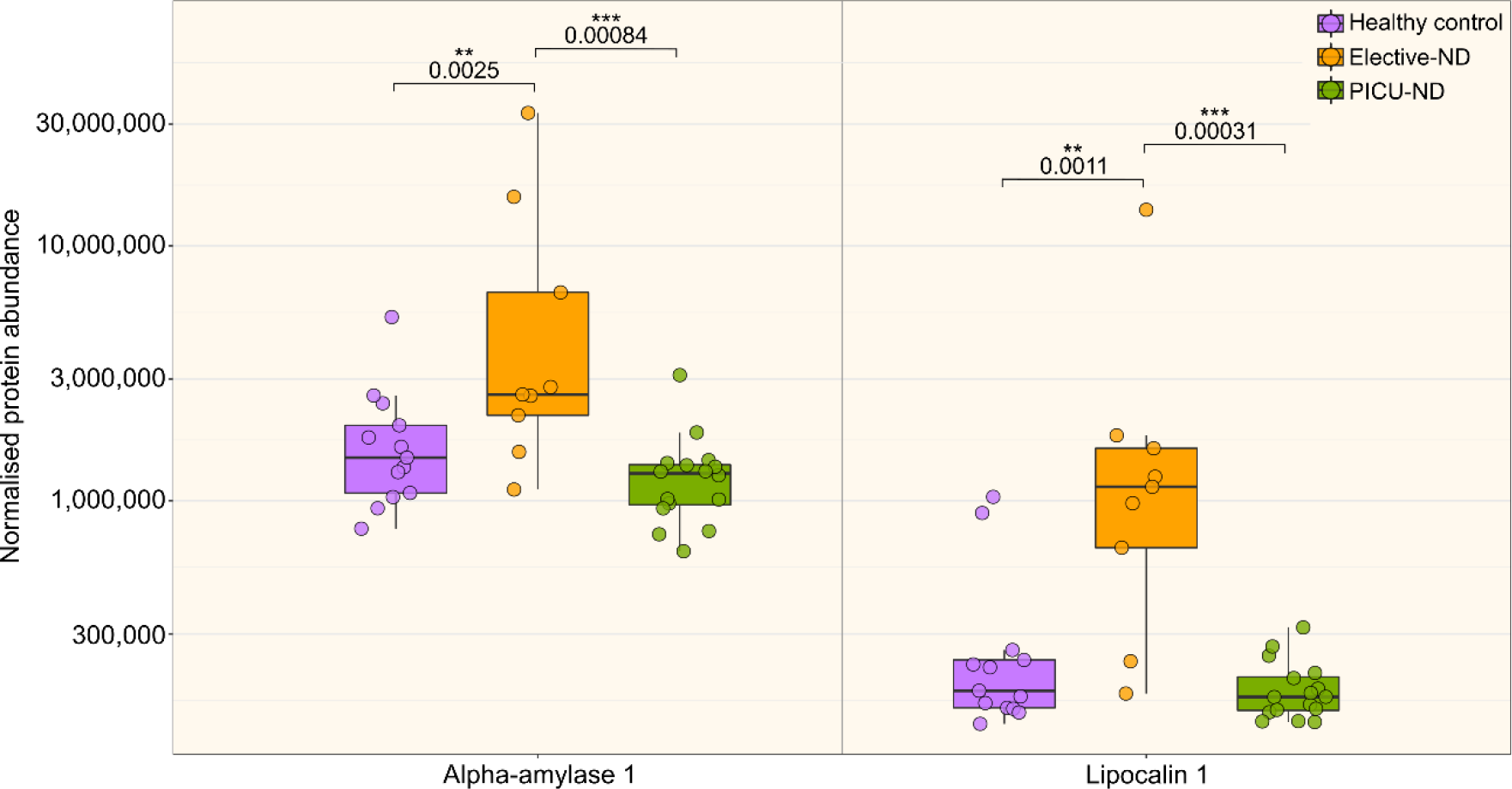
Expression of candidate aspiration proteins in bronchoalveolar lavage fluid (BALF) from healthy control children and children with severe neurodisability (ND). Two proteins enriched in saliva were identified in BALF samples from healthy control children (purple), children with elective-ND (orange) and PICU-ND (green). Label-free abundances for individual samples are plotted, together with a box and whiskers plot (median, box: 50 percentile limits, whiskers, 25th and 75th percentiles). An asterisk (*) indicates statistical significance (Mann Whitney U p-value < 0.05), two (**) indicate a p-value < 0.01 and three (***) indicate a p-value < 0.001.

In total, 291 proteins were identified in gastric fluid. Of these, four were gastric specific (pepsin A, gastrokine-1, gastric triacylglycerol and gastric intrinsic factor), but none were detected in BALF, either by LC-MS/MS or western blot (data not shown).

## Conclusions

We provide evidence of direct, but not reflux, aspiration in children with severe ND. Saliva specific proteins, alpha-amylase-1 and lipocalin-1 were increased 2.0-fold and 6.6-fold respectively in BALF of elective-ND patients compared to healthy control children. Lower levels of these proteins were identified in BALF from controls suggesting for the first time that small amounts of salivary aspiration in health is likely normal. Low levels were also identified in PICU-ND group. We speculate that these patients experienced some degree of airway protection by being intubated for up to 24 hours prior to BALF sampling.

It is unknown whether alpha-amylase-1 and lipolcalin-1 have pathogenic effects in the respiratory tract. However, we have previously shown that oral commensals are frequently found in lower respiratory tract secretions from children with ND.^4^ It is likely that in some children with ND, saliva and bacteria are continuously aspirated. This may account for their increased respiratory symptoms and potentially why some children respond to prophylactic antibiotics.

No potential biomarkers of reflux aspiration were identified in BALF from children with severe ND at the time of sampling. This does not mean that reflux aspiration does not occur in these children, just that if it had occurred, it wasn’t in the immediate pre-operative period (perhaps not surprising as patients will have been starved before surgery). Equally, potential biomarker abundance could have been below the lower limit of detection for the assay.

## Data Availability

All data produced in the present study are available upon reasonable request to the authors.

---

**Table 1.**
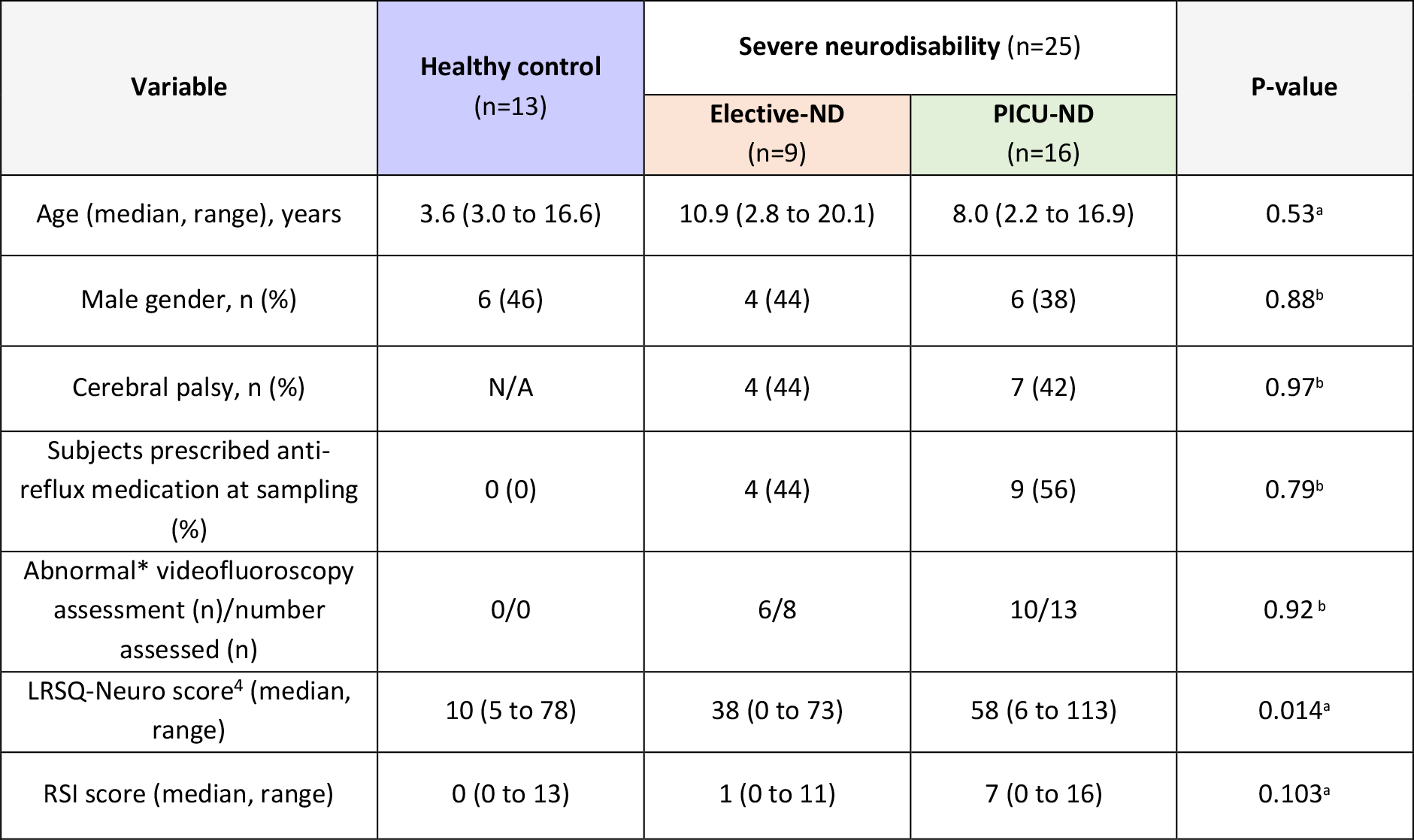
Subject demographics. LRSQ-Neuro; Liverpool Respiratory Symptom Questionnaire – Neuro. RSI; Reflux Symptom Index. *Clear aspiration or transient laryngeal penetration on videofluoroscopy ^a^One-way ANOVA, significance level P < 0.05 ^b^Chi-square test, significance level P < 0.05

